# Understanding Demographic Disparities and Personal Barriers to Self-Care in the UK: Findings from the Living Self-Care Survey Study

**DOI:** 10.1101/2025.06.01.25328757

**Authors:** Austen El-Osta, Sami Altalib, Mahmoud Al Ammouri, Peter Smith

**Author notes:** Corresponding author: (AEO). **Author Contributors:** All authors provided substantial contributions to the conception (PS, AEO), design (AEO, PS, SA, MA), acquisition (SA, AEO) and interpretation (PS, SA, MA, DM, RB, AEO) of study data and approved the final version of the paper. AEO took the lead in planning the study with support from co-authors. SA and MA carried out the data analysis with support from AEO. AEO is the guarantor.

## Abstract

**Background:** Understanding how people manage everyday health challenges is vital for developing self-care policies that are inclusive, equitable and effective. Little is known about how demographic characteristics shape self-care confidence, symptom management strategies and health-seeking behaviours across the UK adult population.

**Objective:** The aim of this study was to examine the demographic predictors of self-care engagement among UK adults, focusing on self-care confidence, health information-seeking behaviours, symptom management strategies and perceived barriers to self-care.

**Methods:** A cross-sectional online survey collected data from 3,255 UK adults, including a subset of health and care professionals (HCPs) to assess self-care knowledge, confidence, practices and health information accessibility. Ordinal logistic regression was used to identify demographic predictors of self-care engagement.

**Results:** Regression analyses highlighted marked demographic disparities in self-care engagement. Older adults (65+) were significantly more confident in their self-care knowledge (aOR=3.22) and healthy lifestyle behaviours (aOR=2.96) yet were also more likely to seek health information. Males reported lower self-care confidence than females (aOR=0.79). Black British participants were more confident in self-care knowledge (aOR=1.99), but along with Asian British individuals, were significantly less likely to seek health information (aORs=0.56 and 0.70 respectively). People living with disabilities (aOR=0.69) and long-term conditions (aOR=0.76) reported lower confidence across self-care domains. HCPs consistently reported higher self-care confidence (aOR=1.51, p=0.006) and health behaviour engagement (aOR=1.66). Financial constraints (53%), lack of time (47%) and low self-efficacy (22%) emerged as key barriers to self-care, alongside low use of pharmacists (1.3%) and digital resources (9.7%).

**Conclusion:** This study highlights demographic disparities in self-care confidence, information-seeking behaviours and barriers to engagement, urging the need for tailored self-care interventions. Future research should explore interventions to improve health literacy, enhance pharmacist-led self-care support and promote equitable access to digital health resources are essential for optimizing self-care engagement across diverse population groups.

## Introduction

There are over 130 definitions of self-care in the academic literature(1), including five definitions from the World Health Organization (WHO). In simple terms, self-care is our ability to maintain our health and wellbeing and to prevent, delay or change the trajectory of common lifestyle diseases(2). It is increasingly recognized as a cornerstone of sustainable health and care systems, particularly in light of growing health and care demands, workforce shortages and the rising burden of chronic diseases(3, 4). Effective self-care practices can reduce pressure on health and care services and empowers individuals to take greater responsibility for their health and wellbeing(5, 6). Despite these recognized benefits, self-care engagement is neither uniform nor equitable across populations, with disparities in self-care confidence, health literacy, access to information and encouragement from health and care professionals (HCPs) contributing to variable adoption and effectiveness of self-care strategies(7–9).

Understanding how individuals engage with and navigate self-care practices is crucial for designing targeted public health interventions that enhance self-care literacy, improve health outcomes and support health and care system sustainability. Previous research, including from the Living Self-Care Survey Study by Smith et al., 2025 examined the public-health and care professional interface, revealing significant demographic disparities in self-care confidence, health-seeking behaviours and engagement with professional guidance(8, 9).

Despite increasing policy emphasis on self-care (3) there remains a limited empirical understanding of how demographic differences influence self-care behaviours and health-seeking practices. One critical gap relates to self-care confidence, which varies across age, gender and ethnicity, yet the underlying factors shaping these differences remain poorly understood(10–12). While older adults report lower self-care confidence, they are simultaneously more likely to seek health information, suggesting potential barriers to translating knowledge into effective self-care actions particularly for people living with multiple long-term conditions(13). Conversely, some ethnic groups exhibit higher confidence but lower engagement with formal health resources (14, 15) raising questions about trust in health and care systems, cultural perceptions of self-care and alternative health-seeking behaviours.

Another pressing issue is health literacy and information-seeking behaviours, which are fundamental to effective self-care but remain highly variable across demographic groups(16–18). The increasing reliance on digital health resources as a means of promoting self-care has created new challenges related to accessibility, digital literacy and the credibility of online health information. Individuals with lower health literacy or limited digital access may struggle to interpret and apply self-care guidance effectively, leading to misinformed decision-making, increased dependency on health and care services or disengagement from self-care practices altogether(19, 20). Understanding how different population groups access, evaluate and trust health information is critical in the context of the ‘symptom iceberg’, alluding to the silent majority of symptoms managed outside formal health and care settings.

The aim of this study was to build on the initial findings from the Living Self-Care Survey Study (Smith et al, 2025) by exploring the demographic predictors, personal barriers and behavioural patterns that influence self-care engagement among UK adults. Specifically, we examined how factors such as age, gender, ethnicity, disability, long-term conditions and professional background affect individuals’ confidence in practicing self-care, their likelihood of seeking health information and their preferred symptom management strategies. We also sought to identify the most reported personal and structural barriers that hinder self-care, including financial limitations, time constraints and lack of access to trustworthy information or professional support.

## Methods

### Study design and data collection

This study is part of the Living Self-Care Survey (LSCS) Study, a cross-sectional, UK-wide online survey investigating self-care confidence, behaviours and health information-seeking practices. The survey was conducted between 1 January and 30 September 2024 (9 months) using the Imperial Qualtrics XM online platform, with participants recruited through health and care networks and social media data collected were stored on the Imperial College London secure database and only the team researchers could access the survey results. All responses were pseudo-anonymised to ensure confidentiality by assigning each respondent a unique study ID. Only the participants’ demographic data (age in years, gender, ethnicity, employment type, highest education level and region of residence) were recorded. Respondents were able to refrain from providing an answer by selecting ‘prefer not to say’).

Full methodological details, including survey development, participant recruitment and data collection procedures, have been previously described in Smith et al., 2025.

### Study participants

The study sample consists of 3,255 community-dwelling UK adults, including a subset of health and care professionals, who completed the LSCS survey. Inclusion and exclusion criteria, survey structure and ethical approvals were detailed in Smith et al., 2025.

### Measures and variables

This study examines self-care engagement through five key domains: (i) Demographic predictors of self-care knowledge, confidence and information accessibility (age, gender, ethnicity, education and HCP status), (ii) Confidence in leading a healthy lifestyle, including engagement with preventive health behaviours, (iii) Health-seeking behaviours and symptom self-management for common conditions, (iv) Personal barriers to self-care, such as financial constraints, time limitations and lack of support and (v) Influence of HCP encouragement on self-care adoption across demographic subgroups.

### Statistical analysis

Survey responses were analysed using descriptive statistics and ordinal logistic regression models to identify demographic predictors of self-care engagement. Adjusted regression models controlled for age, gender, ethnicity, education, employment status, disability, long-term conditions and HCP status. The results were reported using odds ratios (OR) and adjusted odds ratios (aOR) with 95% confidence intervals (CI). Statistical significance was set at p <0.05. The reference categories for categorical variables were consistently defined across analyses (e.g., age 18-24, female gender, White ethnicity).

All analyses were conducted using STATA version 17 (StataCorp LP, College Station, TX, USA). The Checklist for Reporting Results of Internet E-Surveys (CHERRIES) was used to guide reporting(21).

### Ethics

The study was given ethical approval by Imperial College Research Ethics Committee (ICREC #6979141). Participants consented to take part in the survey by completing relevant tick box consent items at the start of the survey.

### Patient and Public Involvement

No patients were involved.

## Results

### Demographic profile of respondents

The survey included 3,255 respondents, of whom the majority were aged between 25-34 years (22.9%) and 35-44 years (22.4%). Participant characteristics are shown in **table 1**. The demographic characteristics of the study sample are itemised in S1Table.

**Table 1:** Actions Taken for Symptoms Experienced in Previous Two Weeks (N = 3,255)

| Symptom | No. | % of N experiencing symptoms | Did nothing/ waited | Used the Internet | Discussed with family & friends | Checked the NHS site | Called 111 | Took over the counter medicines | Consulted a nurse | Consulted a pharmacist | Consulted a GP | Consulted complementary therapist | Took a prescribed medicine |
| --- | --- | --- | --- | --- | --- | --- | --- | --- | --- | --- | --- | --- | --- |
| Headaches | 1,610 | 49.5% | 403 (25.0%) | 72 (4.5%) | 88 (5.5%) | 35 (2.2%) | 8 (0.0%) | 1,157 (71.9%) | 8 (0.5%) | 11 (0.68%) | 13 (0.8%) | 4 (0.3%) | 61 (3.8%) |
| Back pain | 1,077 | 33.1% | 460 (42.7%) | 106 (9.4%) | 98 (9.1%) | 45 (4.2%) | 4 (0.4%) | 441 (40.9%) | 12 (1.1%) | 11 (1.0%) | 36 (3.3%) | 24 (2.2%) | 85 (79.0%) |
| Sore throat | 427 | 13.1% | 186 (43.6%) | 25 (5.9%) | 25 (5.9%) | 12 (2.8%) | 5 (1.2%) | 215 (50.3%) | 4 (0.9%) | 10 (2.4%) | 8 (1.9%) | 0 (0.0%) | 9 (2.1%) |
| Indigestion / Heartburn | 584 | 17.9% | 148 (25.3%) | 40 (6.0%) | 28 (4.8%) | 12 (2.1%) | 2 (0.3%) | 343 (58.7%) | 2 (0.3%) | 10 (1.7%) | 11 (1.9%) | 4 (0.7%) | 77 (13.2%) |
| Cough | 455 | 14.0% | 228 (50.1%) | 30 (6.6%) | 30 (6.6%) | 14 (3.1%) | 1 (0.2%) | 168 (36.9%) | 9 (2.0%) | 13 (2.9%) | 17 (3.7%) | 0 (0.0%) | 18 (3.9%) |
| Cold or flu symptoms | 412 | 12.7% | 153 (37.1%) | 20 (4.9%) | 32 (7.7%) | 16 (3.8%) | 1 (0.2%) | 233 (56.5%) | 7 (1.7%) | 10 (2.4%) | 10 (2.4%) | 0 (0.0%) | 11 (2.7%) |
| Joint pain | 881 | 27.1% | 467 (53.0%) | 40 (4.5%) | 35 (4.0%) | 25 (2.8%) | 6 (0.7%) | 373 (42.3%) | 3 (0.3%) | 5 (0.6%) | 12 (1.4%) | 2 (0.2%) | 19 (2.2%) |
| Covid | 53 | 1.6% | 10 (18.9%) | 8 (15.1%) | 9 (17.0%) | 13 (24.5%) | 1 (1.9%) | 30 (56.6%) | 2 (3.8%) | 1 (1.9%) | 4 (7.6%) | 1 (1.9%) | 4 (7.5%) |
| Diarrhoea | 397 | 12.2% | 230 (57.9%) | 26 (6.6%) | 12 (3.0%) | 13 (3.2%) | 5 (1.3%) | 116 (29.2%) | 5 (1.3%) | 3 (0.8%) | 13 (3.3%) | 1 (0.3%) | 18 (4.5%) |
| Constipation | 319 | 9.8% | 163 (51.1%) | 43 (13.5%) | 28 (8.8%) | 10 (3.1%) | 1 (0.3%) | 110 (34.5%) | 0 (0%) | 7 (2.2%) | 8 (2.5%) | 0 (0.0%) | 17 (5.3%) |
| All<br>Symptoms | 6,215 | - | 2,448 | 410 | 385 | 195 | 34 | 3,186 | 52 | 81 | 132 | 36 | 319 |
| % of all<br>symptoms | - | - | 39.4 | 6.6 | 6.2 | 3.1 | 0.5 | 51.3 | 0.8 | 1.3 | 2.1 | 0.6 | 5.1 |

### Main survey findings

The results of the main survey are shown in S2 File.

#### Prevalence of common symptoms and self-care responses

A significant proportion of respondents reported experiencing common symptoms in the two weeks prior to the survey, with headaches (49.5%), back pain (33.1%) and joint pain (27.1%) being the most frequently reported. Other common symptoms included sore throat (13.1%), indigestion/heartburn (17.9%), cough (14.0%) and cold or flu symptoms (12.7%). Gastrointestinal symptoms such as diarrhoea (12.2%) and constipation (9.8%) were also noted, while COVID-19 symptoms were reported by 1.6% of respondents.

#### Symptom management strategies

Most participants managed their symptoms independently, with self-care behaviours varying by symptom type. Headaches (71.9%), indigestion/heartburn (58.7%) and cold or flu symptoms (56.5%) were primarily treated with OTC medications, while other conditions such as back pain (42.7%) and joint pain (53.0%) were more often left untreated, with individuals opting to wait for symptoms to resolve; Table 1, S2 Table. Reliance on health and care professionals was minimal across all symptoms, with 2.1% of respondents consulting a GP, except for COVID-19 symptoms (7.6%) and back pain (3.3%). Pharmacist consultations were less common at 1.3% for all symptoms.

#### Patterns of self-care and health and care utilization

The preference for OTC medication over seeking professional care suggests confidence in self-managing minor ailments. However, the low engagement with pharmacists (1.3%) and digital health resources (9.7%), including only 3.1% that checked the NHS website, and 6.5% that sued the internet. Less than one percent of respondents contacted NHS 111, suggesting that urgent care services were not a primary choice for symptom management.

Findings indicate a strong inclination toward self-management, with waiting and OTC medications being the predominant strategies. When faced with common minor ailments such as headaches, back pain, or sore throats, the two most frequently adopted self-care strategies were taking OTC medications and waiting or doing nothing to see if symptoms improve. For instance, among those who experienced headaches (n=1,610), 71.9% used OTC medicines, and 25.0% chose to wait without seeking further advice or treatment. Similarly, for sore throats (n=427), 50.4% took OTC remedies and 43.6% opted to wait. These patterns were consistent across various symptoms, including indigestion (58.7% used OTC meds) and cold/flu symptoms (56.6% used OTC meds, 37.1% waited). This high reliance on OTC products and passive monitoring highlights a cultural and behavioural tendency toward independent health management, especially for low-acuity conditions. It is further supported by self-reported data indicating that 46.5% “always” and 43.5% “usually” manage their health and common conditions themselves, with 56.3% agreeing and 37.9% strongly agreeing that they actively seek health information when experiencing symptoms. Importantly, confidence levels in managing health were high, with over 91% of participants reporting that they felt “fairly” or “very” confident in their knowledge and understanding to manage common illnesses. This self-efficacy was most pronounced in older adults (aOR=2.63, 95% CI: 1.84–3.78, p<0.001 for ages 55-64), suggesting that age may enhance perceived competence in self-care

#### Personal barriers to self-care

The most frequently cited barrier was financial limitations, with 53.2% of respondents (n=1,731) indicating that a lack of money prevented them from staying healthy or taking care of their common self-treatable or long-term health conditions. Similarly, time constraints were a major concern, with 46.7% (n=1,520). A lack of confidence (22.2%) and insufficient support from HCPs (GPs/practice nurses: 20.1%; consultants/specialists: 16.4%) emerged as significant impediments. Nearly a third (18.1%, n=589) cited a lack of knowledge about health and health and care issues, while 15.9% (n=519) reported difficulties accessing appropriate treatment information. Over a quarter (17.3%, n=563) indicated that a lack of trustworthy information limited their ability to engage in self-care, whereas 8.2% (n=267) reported difficulty in understanding available health information highlighting a potential gap in the accessibility and clarity of self-care resources. Other commonly reported barriers included a lack of home monitoring equipment (18.9%), a lack of training or skills (12.1%) and low personal interest in self-care (16.0%) indicating that motivational and educational interventions may be necessary to enhance engagement. Only a small proportion of respondents (5.4%) selected “Other,” while 1.5% (n=49) reported being unsure about what prevented them from engaging in self-care.

#### Demographic predictors of self-care knowledge, confidence and information accessibility

The ordinal logistic regression analysis presented in Fig 1 and S3 Table identified significant demographic factors influencing confidence in knowledge and understanding of self-care practices. Respondents in the 55-64 age group had more than double the odds of reporting confidence in their self-care knowledge compared to the 18-24 reference group (aOR=2.05, 95% CI: 1.45-2.89, p<0.001). Confidence increased further in the 65+ group (aOR=3.22, CI: 2.06-5.03, p<0.001). Males were less likely to report confidence to self-care compared to females (aOR=0.79, CI: 0.68-0.91, p=0.002). Ethnic differences were also significant; Black British individuals were more likely to report higher confidence than White participants (aOR=1.99, CI: 1.39-2.84, p<0.001), while Asian British individuals were less likely (aOR=0.66, CI: 0.49-0.89, p=0.006).

**Fig 1:** Odds ratios and confidence intervals - knowledge and understanding to practice self-care. Forest plot showing the odds ratios and confidence intervals of factors associated with knowledge and understanding to practice self-care

HCPs reported higher self-care confidence than non-professionals (aOR=1.51, CI: 1.13-2.02, p=0.006) highlighting the role of professional knowledge in self-care practices. Confidence in information accessibility also varied: although 68% found health information easy to access, issues persisted in mental health literacy and treatment evaluation.

### Confidence in leading a healthy lifestyle

Confidence in the ability to lead a healthy lifestyle was influenced by age, ethnicity, education and health status (Fig 2, S4 Table). Older age groups demonstrated greater confidence compared to the 18-24 group, with participants aged 55-64 over twice as likely to report high confidence (aOR=2.11, CI: 1.48-3.00, p<0.001) and those aged 65+ nearly three times as likely (aOR=2.96, CI: 1.87-4.68, p<0.001). Ethnic disparities were also observed; Asian British participants were significantly less likely to report confidence than White participants (aOR=0.46, CI: 0.34-0.63, p<0.001).

**Fig 2:** Odds ratios and confidence intervals - confidence in leading a healthy lifestyle. Forest plot showing the odds ratios and confidence intervals of factors associated with confidence in leading a healthy lifestyle

Participants with secondary education were less likely to feel confident in leading a healthy lifestyle compared to those with primary education (aOR=0.21, CI: 0.05-0.89, p=0.034). Similarly, individuals with a disability or a long-term condition reported lower confidence (disability: aOR=0.58, CI: 0.45-0.74, p<0.001; long-term condition: aOR=0.65, CI: 0.55-0.77, p<0.001). HCPs were notably more confident in their ability to lead a healthy lifestyle (aOR=1.66, CI: 1.24-2.23, p=0.001).

#### Seeking health information for common conditions

The ordinal logistic regression analysis (Fig 3, S5 Table) explored demographic factors influencing the likelihood of seeking health information when experiencing a common condition. Older participants were more likely to seek health information when experiencing common conditions, with individuals aged 55-64 having a 91% higher likelihood than the 18-24 group (aOR=1.91, p<0.001). Females were more proactive in seeking information than males (aOR=0.73, p<0.001), while Black and Asian British individuals were less likely than White participants to seek information (aORs=0.56, 0.70; p<0.05).

**Fig 3:** Odds ratios and confidence intervals of factors associated with finding health information when faced with a common condition. Forest plot showing the odds ratios and confidence intervals of factors associated with finding health information when faced with a common condition

## Discussion

This study builds upon the findings of The Living Self-Care Survey Study (Paper 1 by Smith et al., 2025) shifting the focus toward public engagement with self-care behaviours by examining demographic predictors, symptom management strategies and personal barriers to self-care.

### Self-care confidence and health-seeking behaviours

We found that older adults (65+) were significantly less confident in self-care despite being more likely to seek health information, whereas younger adults exhibited greater confidence but lower engagement with health resources. This aligns with previous research suggesting that while older adults may prioritize health-seeking behaviours, they may lack the necessary health literacy or self-efficacy to act on self-care guidance effectively(22). That men reported lower self-care confidence than women also aligns with existing evidence that women are more likely to engage in proactive health behaviours and seek medical advice(23).

In contrast to earlier studies which often reported low self-care confidence in minority ethnic groups(24, 25), our data reveal a more complex picture. Black and Asian British participants reported higher levels of self-care confidence compared to their White counterparts but paradoxically were less likely to seek health information. This divergence may reflect cultural norms, reliance on community or family advice, or lower trust in formal healthcare channels-a finding echoed in the work of Berkman et al., (26), who emphasised the interplay between culture, trust and health-seeking behaviour.

Additionally, our data suggested that HCPs exhibited significantly greater self-care confidence than the general population, consistent with findings from previous studies on health literacy and professional knowledge (27) translation. However, the influence of HCPs as self-care advocates appears underutilised, with 20% of participants citing a lack of professional encouragement as a barrier. This supports prior calls for HCPs to take a more active role in promoting self-care(28).

### Barriers to self-care and implications for health literacy

The identification of financial constraints (53%) and lack of time (47%) as key barriers highlights the structural challenges that limit self-care engagement, particularly for lower-income and working-age populations, align with existing research highlighting that socioeconomic disparities affect the ability to prioritize preventive health behaviours and access self-care resources. Additionally, a lack of confidence (22%) and limited trust in available health information (17%) suggest that health literacy remains a critical determinant of self-care engagement.

Despite the increasing availability of digital health resources, it was surprising that engagement with online information sources remained low, with fewer than 7% of participants going online or checking the NHS website for symptom management. This suggests that digital exclusion, difficulties in navigating online health content and concerns over misconceptions may hinder the effectiveness of digital self-care interventions(29). Collectively these insights highlight the urgency of embedding health equity principles into national self-care strategies, particularly given the current emphasis on health system sustainability and population resilience.

### Symptom management strategies and health and care utilization

The study highlights a strong inclination toward self-care, with OTC medication use and passive waiting being the dominant management strategies for most symptoms. While this suggests confidence in managing minor ailments, the low rates of engagement with pharmacists and HCPs indicate potential gaps in accessing expert guidance when needed. Given that <2% of participants consulted a pharmacist for symptom management, there is a missed opportunity to leverage pharmacists as accessible self-care advisors as recommended by national self-care policy frameworks.

Additionally, around 2% of respondents sought GP consultations for most symptoms, except for COVID-19 symptoms (8%) and back pain (3%), suggesting that individuals predominantly reserve health and care visits for perceived severe or persistent conditions. While avoiding unnecessary GP visits aligns with self-care promotion goals, there is a need to ensure that individuals can recognize when professional intervention is necessary. Targeted health literacy initiatives could help individuals differentiate between self-treatable conditions and symptoms requiring medical attention, reducing both avoidable consultations and health risks associated with delayed care).

### Symptom iceberg

The findings of this study reinforce and extend existing evidence regarding self-care behaviours, symptom management strategies and barriers to engagement in the UK population. Notably, our results support the foundational work by Elliott et al. (30) who in 2011 described the persistence of the “symptom iceberg” phenomenon where the majority of common symptoms are managed privately rather than presented to primary care (24). Our study, conducted over a decade later, suggests a marked intensification of this pattern, with over 90% of respondents managing symptoms independently, largely due to the use of OTC medicines increasing from 25.0% to 51.3%, and minimal engagement with pharmacists (1.3%) or GPs (2.1%) across most conditions. Compared to Elliott’s estimate of 45% self-care for symptoms such as headaches or sore throats, our findings indicate a significant upward shift, although the underlying drivers of empowerment, digital information access and constrained service availability require further exploration. Comparatively, our findings also suggest a decrease in the ‘tip of the iceberg’, with consultations with any professional reducing from 13.2% to 5.3%. Consultations with GPs reduced from 8.3% to 2.1% and with pharmacists from 1.8% to 1.3%.

### Health services utilisation

These findings also align with contemporary analyses of changing health service utilisation Ladds et al., 2020. (23)note the shift toward remote assessment and digital-first models, particularly following COVID-19, which may partly explain reduced engagement with face-to-face professional advice. However, our findings challenge the assumption that digital uptake is universal; engagement with digital resources (e.g. NHS websites, symptom checkers) remained below 10%, especially among older and lower socioeconomic groups(31, 32). This reflects earlier health literacy research by Nutbeam (24) and Sørensen et al. 2012, which identified persistent gaps in the functional & interactive skills required to engage with digital health tools effectively(27).

### Study implications

This study provides fresh and critical insights into how UK adults manage common health symptoms, highlighting persistent demographic disparities and deeply rooted structural barriers to self-care. The data highlights the continued relevance- and evolution-of the “symptom iceberg” in today’s digitally connected yet unevenly accessible health landscape. These findings have significant implications for health system strategy, public health policy and service design, particularly in the context of preventive care, digital transformation and health equity.

The widespread engagement with self-care behaviours revealed in this study-especially reliance on OTC medications and passive symptom monitoring-suggests that the UK public is already navigating a large proportion of their health concerns outside the formal care system. However, structural and demographic barriers such as financial constraints, lack of time, low confidence and digital exclusion disproportionately limit self-care access and efficacy for key population groups, particularly people living with long-term conditions, disabilities and those from ethnic minority backgrounds. These disparities position self-care not simply as a matter of individual choice but as a structural and social justice issue requiring systemic intervention.

Despite strong policy endorsement of self-care, this study shows that few individuals turn to professionals-particularly pharmacists-for support. Engagement with pharmacists for symptom management remained below 3% across nearly all symptoms. This underutilisation highlights a missed opportunity. Community pharmacies are well-placed to serve as accessible hubs for self-care guidance, but this will require strategic investment in pharmacist training, remuneration for self-care consultations and public education to shift perceptions of pharmacists as self-care enablers.

An important paradox emerging from this study is the mismatch between reported self-care confidence and actual information-seeking behaviour-particularly among older adults and some ethnic minority groups. These findings highlight the limitations of assuming confidence alone predicts behaviour. Health literacy interventions must go beyond general messaging and target the underlying drivers of this disconnect motivational beliefs, information appraisal skills, cultural perceptions of care and trust in available sources. Initiatives should include co-designed content, culturally competent materials and trusted messengers embedded in local communities.

While digital health is increasingly central to self-care promotion, our data show that fewer than 7% of participants consulted trusted online resources like the NHS website during symptom episodes. This low engagement challenges the assumption that digital-first strategies are reaching or resonating with the public. Digital health content must be reimagined for accessibility linguistically, culturally and functionally. This includes plain language, intuitive design, mobile-first interfaces and embedded signposting to real-world support (e.g., community pharmacies, local self-care workshops). National strategies should also fund digital navigation roles and peer educators to improve confidence in using these tools.

The expanded symptom iceberg we observed-with over 90% of common conditions managed independently-raises concern about missed or delayed care, particularly among populations with low self-efficacy or limited access to guidance. Policymakers and public health agencies should promote structured symptom triage tools (digital or paper-based) that guide users through symptom severity, duration and red flags, linking them to the appropriate level of care. These tools should be embedded into public health campaigns and digital portals and integrated within pharmacy and GP reception settings.

Given the variability in self-care confidence and engagement, there is a strong case for incorporating brief self-care readiness assessments into existing patient pathways-such as NHS Health Checks, chronic disease reviews or vaccination appointments. These tools can help clinicians identify individuals who may benefit from targeted support, including health coaching, literacy interventions or peer navigation. A standardised tool, derived from items used in this study, could enable scalable, comparable assessments of self-care capability across settings and populations.

The dominant barriers to self-care identified - money, time, confidence, information and access - span beyond the health sector and demand joined-up action. Government and local authorities should work in partnership with employers, housing providers, education institutions and third-sector organisations to build supportive ecosystems for self-care. Initiatives could include flexible workplace wellbeing schemes, self-care skills in school curricula, social prescribing for digital literacy training and voucher schemes to subsidise self-care essentials like thermometers or OTC medications for low-income households.

With mounting pressures on general practice and urgent care, the findings support the development of a national symptom management strategy that explicitly outlines how common, self-limiting conditions can be safely managed by the public with professional support as needed. This would include clear care pathways, a public-facing symptom reference guide and greater investment in communications that empower people to care for themselves while knowing when- and how-to seek help.

### Limitations

This study has several limitations that were acknowledged in **Smith et al, 2025**. Briefly, the reliance on self-reported data introduces the possibility of recall bias and social desirability effects, particularly in the reporting of health behaviours, confidence and symptom management strategies. Although the survey was anonymous, participants may have over- or under-estimated their engagement with self-care or health information-seeking behaviours. Second, the cross-sectional design limits causal inference. While we identified significant associations between demographic variables and self-care engagement, we cannot determine whether these factors directly influence self-care behaviour or whether unmeasured variables, such as prior health experiences, social support, or mental health status, may mediate these relationships. Third, the study sample was recruited online and may not be fully representative of the UK population. Individuals with limited digital access, lower levels of digital literacy, or those disengaged from online health discourse may have been under-represented. As a result, estimates of engagement with digital health resources and confidence in self-care may be biased toward more digitally literate respondents. Fourth, while our regression models adjusted for a wide range of demographic variables, residual confounding cannot be excluded. For instance, variables such as income, health insurance status (e.g. private vs NHS-only care), caregiving responsibilities, or migration status were not included but may influence self-care capability and behaviour. Finally, although the study explored a range of barriers to self-care, it did not include qualitative data to contextualise individual choices or explore the influence of cultural beliefs, trust in health systems, or lived experiences of marginalisation. Future mixed-methods research is needed to deepen our understanding of the drivers behind observed disparities in self-care engagement.

## Conclusion

This study highlights demographic disparities in self-care engagement, emphasizing the influence of confidence, health literacy, financial constraints and access to reliable health information on self-care behaviours. While self-care was the preferred approach for managing common symptoms, low engagement with pharmacists, digital resources and professional health and care support suggests that further efforts are needed to optimize self-care literacy and accessibility. Addressing structural barriers, enhancing pharmacist roles and improving digital health strategies are essential to ensuring that self-care initiatives effectively reach all population groups. Future research should explore interventions that enhance self-care confidence, reduce inequalities and support sustainable health and care models.

## Data Availability

All relevant data are within the manuscript and its Supporting Information files.

## Declarations

### Data sharing statement

The data that support the findings of this study are contained within S1 File.

### Funding

Financial support was provided as a Quality Improvement Grant from Pfizer to the Self-Care Forum. AEO is in part supported by the National Institute for Health and Care Research (NIHR) Applied Research Collaboration (ARC) Northwest London. The views expressed are those of the authors and not necessarily those of the NHS or the NIHR or the Department of Health and Social Care.

#### Acknowledgments

The authors thank the Self-Care Forum for disseminating the link to the survey

## Twitter

@austenelosta @ImperialSCARU @SamiAltalib

## Supporting Information

**S1 Table: Demographic characteristics of the study sample.doc**

**S2 Table: Managing Common Conditions**

**S3 Table: Factors predicting knowledge and understanding to practice self-care**

**S4 Table: Factors predicting the confidence with the knowledge and understanding to lead a healthy lifestyle**

**S5 Table: Factors predicting ability to find relevant health information when faced with a common condition**

**S1 File: Raw Data.xls**

**S2 File: Survey Findings.doc**

**S1 Fig:**
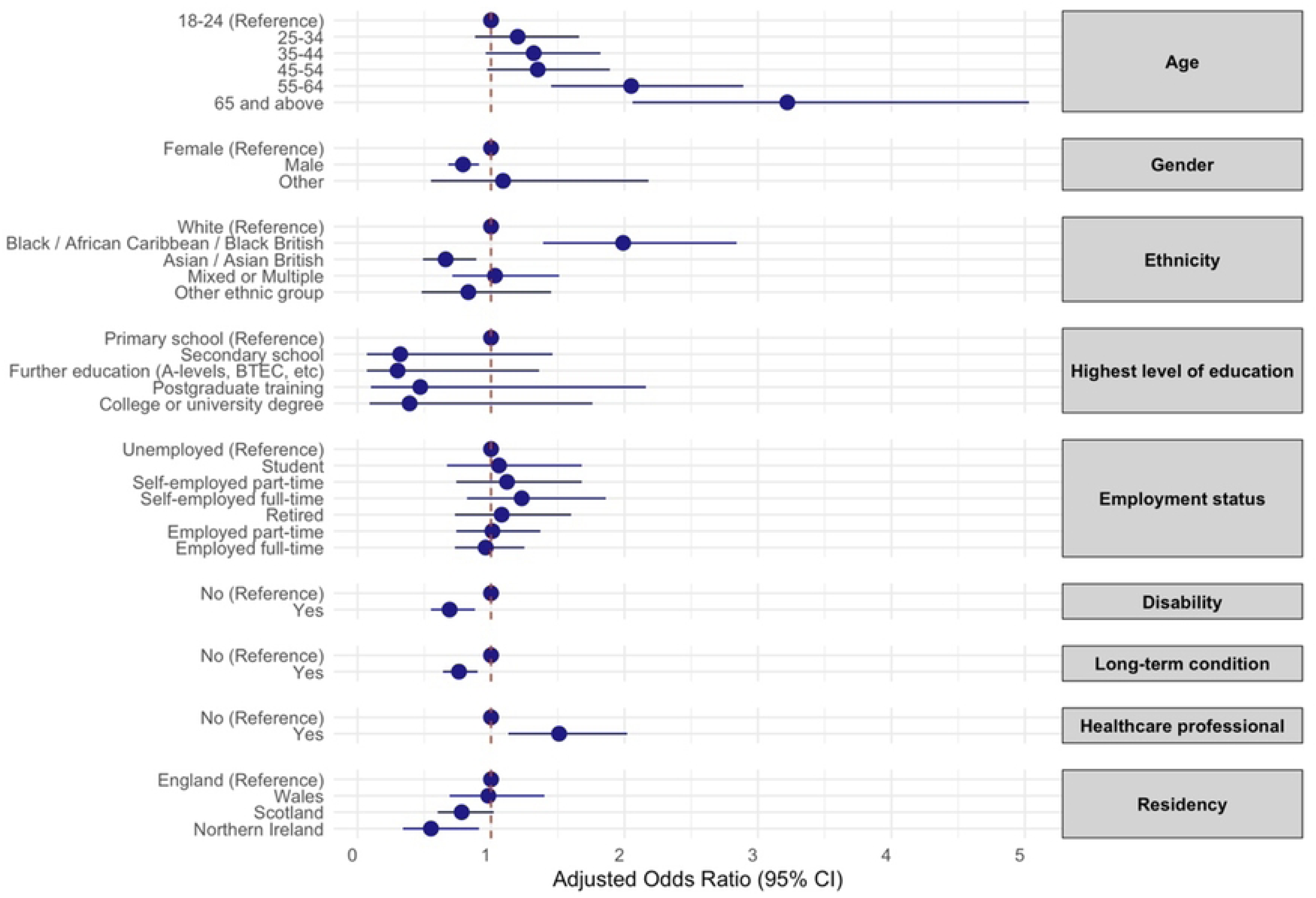
Knowledge and understanding to practice self-care. Forest plot showing the odds ratios and confidence intervals of factors associated with knowledge and understanding to practice self-care.

**S2 Fig:**
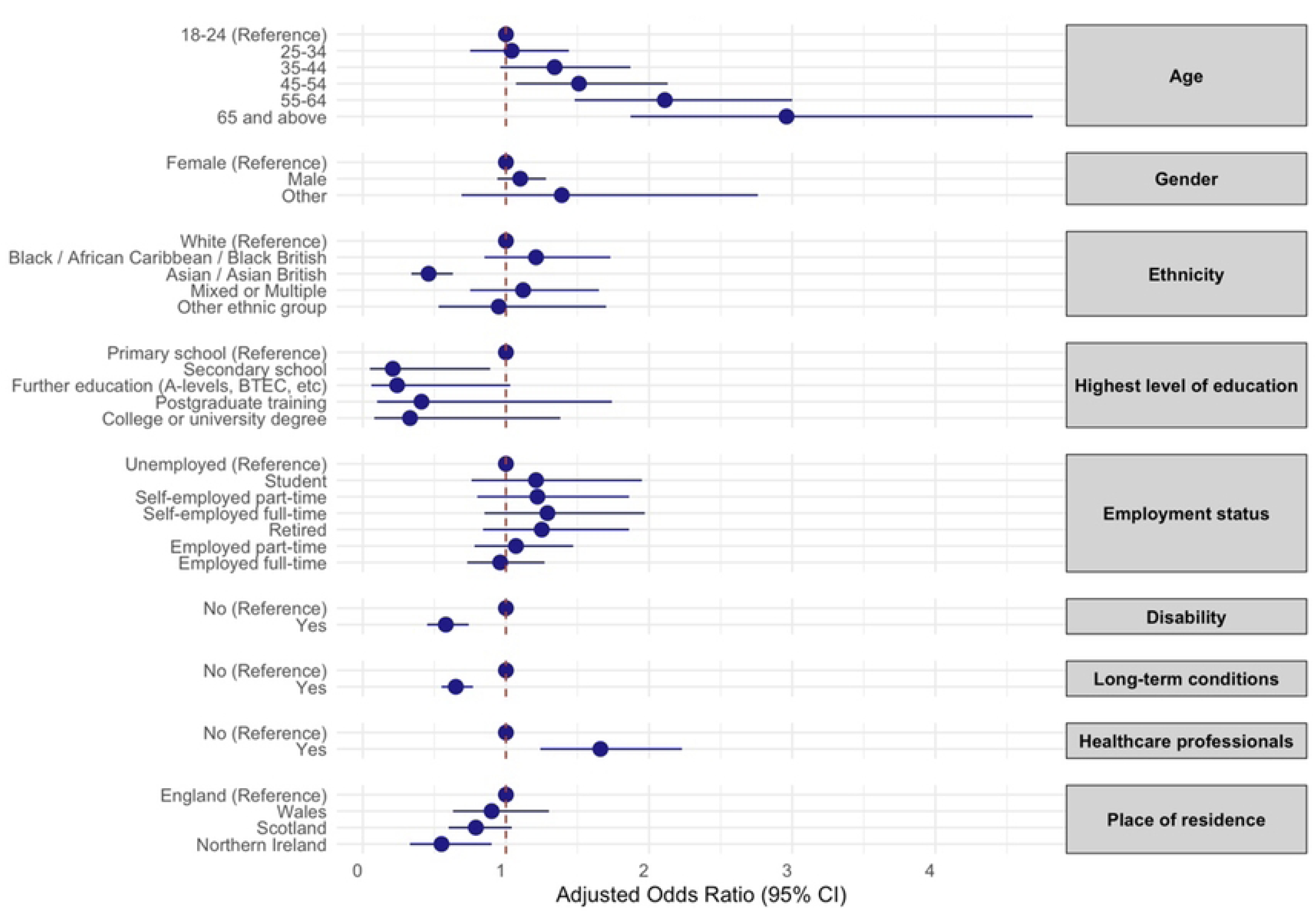
Confidence in leading a healthy lifestyle. Forest plot showing the odds ratios and confidence intervals of factors associated with confidence in leading a healthy lifestyle

**S3 Fig:**
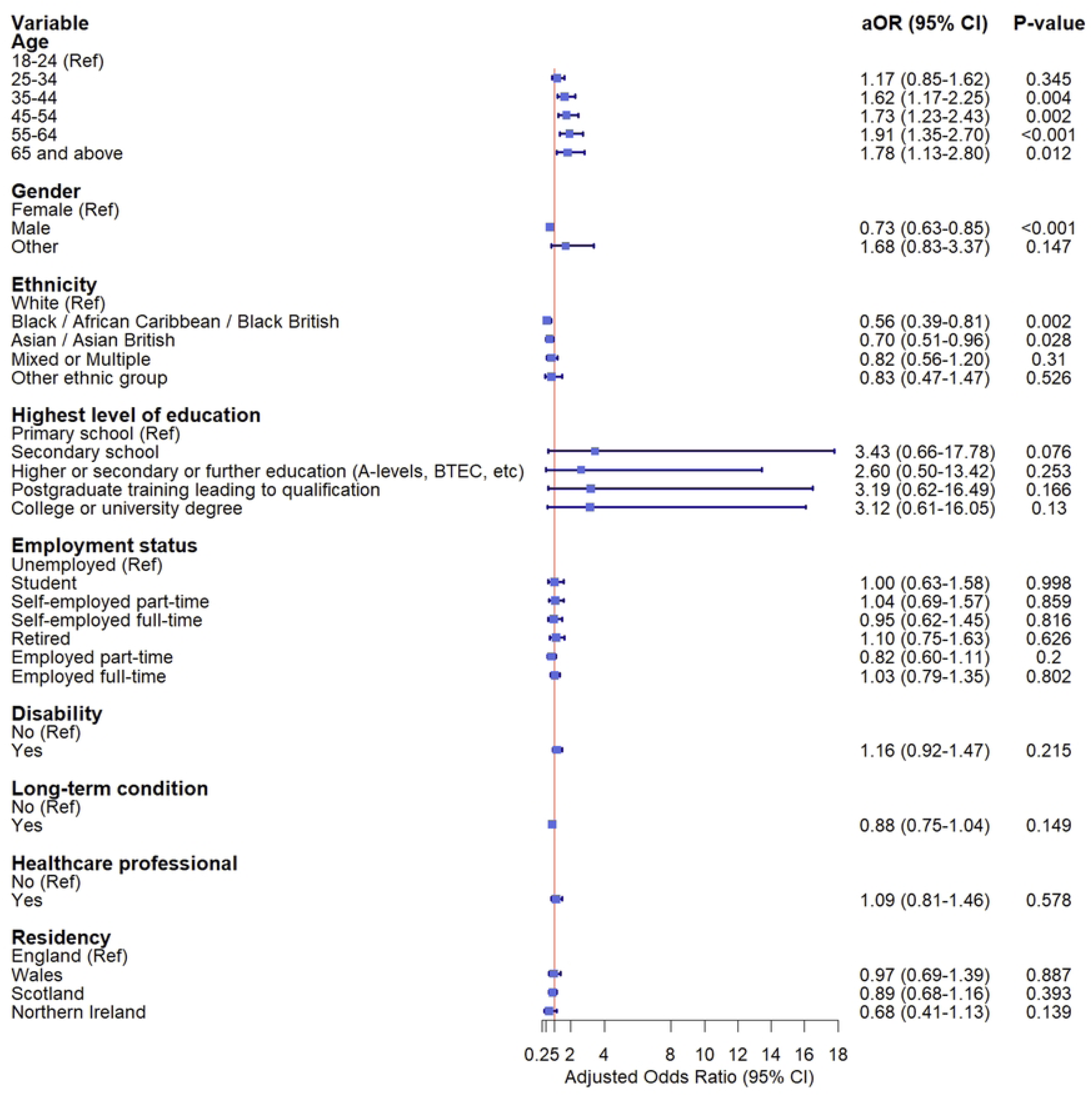
Finding health information when faced with a common condition. Forest plot showing the odds ratios and confidence intervals of factors associated with finding health information when faced with a common condition

## Notes

### Competing Interest Statement

The authors have declared no competing interest.

### Funding Statement

Yes

### Author Declarations

The study was reviewed and given ethical approval by Imperial College Research Ethics Committee (ICREC #6979141). Participants consented to take part in the survey.

